# Light at night and modeled circadian disruption predict higher risk of mortality: A prospective study in >88,000 participants

**DOI:** 10.1101/2023.09.08.23295231

**Authors:** Daniel P. Windred, Angus C. Burns, Jacqueline M. Lane, Patrick Olivier, Martin K. Rutter, Richa Saxena, Andrew J. K. Phillips, Sean W. Cain

**Affiliations:** Turner Institute for Brain and Mental Health, School of Psychological Sciences, Faculty of Medicine, Nursing and Health Sciences, Monash University, Melbourne, VIC, Australia; Center for Genomic Medicine, Massachusetts General Hospital, Boston, MA, USA; Department of Anesthesia, Critical Care and Pain Medicine, Massachusetts General Hospital and Harvard Medical School, Boston, MA, USA; Division of Sleep and Circadian Disorders, Brigham and Women’s Hospital, Boston, MA, USA; Action Lab, Department of Human-Centred Computing, Faculty of Information Technology, Monash University, Melbourne, VIC, Australia; Centre for Biological Timing, Division of Endocrinology, Diabetes & Gastroenterology, School of Medical Sciences, Faculty of Biology, Medicine and Health, Manchester Academic Health Science Centre, University of Manchester, Manchester, UK; Diabetes, Endocrinology and Metabolism Centre, NIHR Manchester Biomedical Research Centre, Manchester University NHS Foundation Trust, Manchester, UK

**Author notes:** **Corresponding Author:** Sean W. Cain. Authors contributed equally to this manuscript (Andrew J. K. Phillips, Sean W. Cain).

## Abstract

**Importance:** Light at night disrupts human circadian rhythms, which are critical for maintaining optimal health. Circadian disruption accompanies poor health outcomes that precede premature mortality, including cardiometabolic diseases. However, links between personal night light exposure and premature mortality risk have not been established.

**Objective:** To characterize the association of light at night with all-cause and cardiometabolic mortality risks and to understand the role of circadian disruption in these associations by applying a computational model of the response of the human circadian pacemaker to light.

**Design:** Prospective cohort study.

**Setting:** United Kingdom.

**Participants:** UK Biobank cohort, N=88,904, aged 62.4±7.8 years, 57% female.

**Exposure:** Participants wore activity tracking watches with light sensors for one week between 2013-2016. Twenty-four-hour light exposure profiles were extracted for each participant, and day-time and night-time hours were defined by factor analysis. A validated mathematical model of the human circadian pacemaker was applied to model circadian amplitude and phase from weekly light data.

**Main Outcome:** Cause-specific mortality (National Health Service) recorded in 2,605 participants across a mean (±SD) follow-up period of 6.31±0.83 years after light/activity tracking.

**Results:** Risk of all-cause mortality was higher in participants in the 90^th^-100^th^ percentiles of night-light exposure (HR[95%CI]=1.30[1.15-1.48]), and for those between the 70^th^-90^th^ percentiles (HR=1.16[1.04-1.28]), compared to the darkest 50%. Participants in the 90^th^-100^th^ percentiles of night-light exposure also had higher risk of cardiometabolic mortality (HR=1.41[1.07-1.85]). Higher circadian amplitude predicted lower risks of all-cause mortality (HR = 0.94[0.91-0.97] per SD) and cardiometabolic mortality (HR=0.90[0.83-0.96]), and circadian phase that deviated from the group average predicted higher risks of all-cause mortality (HR=1.33[1.17-1.51]) and cardiometabolic mortality (HR=1.48[1.12-1.97]). These findings were robust to adjustment for age, sex, ethnicity, and sociodemographic and lifestyle factors.

**Conclusions and Relevance:** Minimizing exposure to light at night and keeping regular light-dark patterns that enhance circadian rhythms may promote cardiometabolic health and longevity.

**Key Points:** *Question:* Is light exposure at night associated with risk of premature mortality?

*Findings:* Exposure to brighter light at night, recorded with personal light sensors in >88,000 participants, was associated with higher risk of mortality across a subsequent 6-year period. Computational modeling indicated that disrupted circadian rhythms may explain this higher mortality risk.

*Meaning:* Avoiding light at night may be a cost-effective and accessible recommendation for promoting health and longevity.

## Introduction

Circadian rhythm disruption accompanies a wide range of adverse health outcomes^1–3^ that contribute to premature mortality. Light exposure at night disrupts circadian rhythms by shifting the timing (phase-shift) and weakening the signal (amplitude suppression) of the central circadian pacemaker in the hypothalamus^4–6^, which orchestrates circadian rhythms throughout the body.^7,8^ Experimental exposure to light at night causes premature mortality in animal models.^9,10^ Human populations who are more likely to be exposed to light at night, including rotating shift-workers,^11^ evening types,^12^ and those with fragmented activity patterns^13,14^ have higher risks of premature mortality. Furthermore, population-scale studies have linked outdoor light at night with higher risk of all-cause mortality^15^ and coronary heart disease,^16^ using satellite data. However, the relationship between objective individual-level light exposure patterns and risk of all-cause mortality in humans has not been investigated.

Circadian disruption leads to cardiometabolic dysfunction and morbidity, which increase mortality risk.^17^ Experimental disruption of circadian rhythms alters blood glucose, insulin, cortisol, leptin, arterial pressure, and energy expenditure.^18^ Myocardial infarction, stroke, hypertension, diabetes, and metabolic syndrome have higher incidence in rotating shift-workers.^19–22^ Cardiovascular risk factors, metabolic syndrome, and high BMI are also more often observed in evening types.^18^ In animal models, disruption of circadian rhythms with light produces profound cardiovascular disease, causing premature death due to cardiomyopathy, extensive fibrosis, and severely impaired contractility.^23^ However, no large-scale study has examined associations of individual- level light exposure with risk for premature mortality by cardiometabolic causes.

We characterized the association of light at night with all-cause and cardiometabolic mortality risk in ∼87,000 UK Biobank participants, using 14 million hours of data from wrist-worn light sensors. We also examined the association of circadian disruption with mortality using a validated computational model of the human circadian system, which allowed us to estimate an individual’s circadian phase and amplitude.

## Methods

### Overview

Approximately 502,000 UK Biobank participants aged between 40-69 years were recruited between 2006- 2010.^24^ From this cohort, 103,669 participants wore Axivity AX3 devices (Axivity, Newcastle upon Tyne, UK) on their dominant wrist for 7 days under free-living conditions (2013-2016). Devices were distributed and returned by post. Light and accelerometer data were logged at 100Hz. Devices contained an APDS9007 silicon photodiode light sensor that responded to a spectral range similar to the human eye (peak sensitivity wavelength of 560nm). We tested a sample of Axivity AX3 devices under reference lighting conditions, confirming an approximately linear response to illuminance between 0-5500 lx. See Supplementary S1, S4, S5 and S10 for additional detail on protocol and device testing.

Mortality data were received by the UK Biobank from NHS Digital (England) and NHS Central Register (Scotland). Records included date of death and primary cause of death, diagnosed according to the ICD-10. Death records between June 2013 and March 2021 were included in analyses.

Cardiometabolic mortality was defined as any cause of death corresponding to ICD-10 diseases of the circulatory system, or endocrine and metabolic diseases. Predominant circulatory causes of death were ischemic heart disease (I20-I25), cerebrovascular diseases (I60-I69), other heart disease (I30-I52), diseases of the arteries, arterioles, and capillaries (I70-I79), and hypertensive diseases (I10-I15). Predominant endocrine and metabolic causes of death were diabetes mellitus (E10-E14), metabolic disorders (E70-E90), and obesity (E65-E68).

The UK Biobank has ethical approval from the North West Multi-centre Research Ethics Committee (https://www.ukbiobank.ac.uk/learn-more-about-uk-biobank/about-us/ethics).

### Light exposure profiles

Light data were cleaned based on accelerometer data to ensure light recordings corresponded to when devices were on-wrist. Device non-wear was determined by GGIR, a validated package for estimating sleep-wake state from accelerometer data, as reported previously.^25–27^ Participants had a median (IQR) of 6.90 (5.95-6.96) days of light data remaining after exclusion of epochs coinciding with non-wear. Participants with no valid days detected by GGIR were excluded, due to non-wear or data corruption (8,004 of 103,669).

Daily profiles of light exposure were extracted for each participant by grouping weekly light data into forty-eight half-hour clock time intervals across 24 h (e.g., 00:00 to 00:30 across 7 days). We excluded participants with low quality light data reflecting device malfunction, or insufficient data in any of the forty-eight half-hour intervals (<60 of 210 possible minutes of cumulative light data across the week; 6,761 of 95,665 participants excluded). Daily light profiles representing all 24 h clock times remained in 88,904 participants. Factor analysis was applied to daily profiles, extracting a two-factor structure of day light (07:30-20:30) and night light (00:30-06:00; see Supplementary Appendix S10).

### Covariates

Physical activity was included as the acceleration average across each weekly recording, as derived in previous work.^28^ Additional covariates were collected between 2006-2010, including self-reported ethnic background, employment status, yearly household income, Townsend Deprivation Index, weekly social activities, frequency of social visits, smoking status, urbanicity, and rotating shift-work status. Cardiometabolic risk factors included BMI, hypertension, cholesterol ratio (total cholesterol/HDL), diabetes diagnosis, and history of vascular conditions. Sleep duration was calculated using GGIR, a validated package for estimating sleep-wake state from accelerometer data, as reported previously.^25–27^ See Supplementary Appendix S2, S3, and S10 for detailed descriptions of covariates.

### Statistical analysis

Light data were split into four percentile groupings: 0-50% (referent group), 50-70%, 70-90%, and 90-100%. The 0-50^th^ percentiles were grouped due to minimal variability in their average light intensity at night, and this group were hypothesized to have the lowest risk of mortality. Hazard ratios for all-cause and cause-specific mortality were estimated using Cox proportional hazards models and competing-risks proportional sub-hazards models.^29^ Time since light/activity recording was used as the timescale and all models were adjusted for participant age. Relationships between light exposure and mortality were assessed using two approaches: (i) models including night and day light factors together, to assess night light/mortality relationships while adjusting for day light exposure, and (ii) models assessing the time-of-day relationship between light exposure and mortality, consisting of 48 models corresponding to half-hour clock time intervals, adjusted for multiple comparisons using a Bonferroni correction (p<.001). Two hierarchical model levels were implemented: Model 1 (minimally-adjusted) included age, sex, and ethnicity, and Model 2 (fully-adjusted) was additionally adjusted for physical activity, employment, income, deprivation, social activities, social visits, smoking status, urbanicity, and shift work status.

Secondary analyses included: (i) an assessment of the temporal stability of light exposure patterns in 2,998 participants with up to four repeated light/activity recordings each, across seasons, (ii) addition of baseline cardiometabolic health factors including diabetes diagnosis, vascular events, BMI, hypertension, and cholesterol ratio to fully-adjusted models, and (iii) assessment of the mediating role of night light in the relationship between short sleep duration and mortality risk (see Supplementary Appendix S7-9, and S11).

### Circadian rhythm modeling

To model circadian amplitude and phase, weekly light data were input to a mathematical model^30^ that approximates the response of human photoreceptors (represented by a dynamic stimulus processor) and the central circadian pacemaker (represented by a limit cycle oscillator) to light exposure. This model has been applied in a wide range of populations and is the best existing method for predicting the state of the human circadian clock from light data.^31^ In the model, light modulates pacemaker phase and amplitude in a time- and state- dependent manner. Light near the middle of the biological night suppresses amplitude, while light in the early and late biological night delays and advances phase, respectively. See Supplementary S10 for model equations and implementation. Amplitude was calculated at each epoch. Mean, minimum, and maximum amplitudes were calculated over this time series (approximately 7 days) for each participant. Phase (predicted time of core body temperature minimum) was calculated for each ∼24 h cycle. Mean and standard deviation of phase across the week were calculated for each participant. Amplitude metrics and phase variability were z-scored and included as continuous predictors of all-cause, cardiometabolic, and non-cardiometabolic mortality across two levels of Cox proportional hazards and sub-hazards models as described above. Mean phase was split into quintiles to account for the circular nature of the data. The 40-60^th^ percentile group was centered at participant’s circular mean phase (03:50) and was used as a referent group in Cox proportional hazards models.

## Results

### Descriptive statistics

Our final analyses included 88,904 participants with complete daily light profiles. Mean follow up period was 6.31±0.83 years between light/activity recording and study endpoint (21^st^ March, 2021), and total follow-up period was 7.8 years. All-cause mortality rate was 4.64 deaths per 1000 person-years, including 2,605 all-cause and 539 cardiometabolic deaths. Participants were 62.4±7.8 years, 57.0% female, 97.0% white ethnicity, 62.1% employed, 8.0% shift-workers, and had median income range £31,000-51,999, Townsend deprivation score of - 1.76±2.80, ≥1 weekly social activities experienced by 72.8%, social visits most commonly experienced weekly (36.3%), 6.8% current and 36.1% previous smokers, 84.1% from an urban postcode, and average physical activity of 28.2±8.1 milli-g across weekly recordings (see Table 1). All-cause, cardiometabolic, and non-cardiometabolic deaths for light exposure percentile groups are detailed in Table 2, alongside approximate ranges of light intensity for each group. The distribution of approximate light intensity across 24 h is provided in Supplementary S4.

**Table 1.** Descriptives statistics for participants grouped according to light intensity percentiles, and split by day and night light.

|  | Day light exposure percentile |  |  |  | Night light exposure percentile |  |  |  |
| --- | --- | --- | --- | --- | --- | --- | --- | --- |
|  | 0-50% | 50-70% | 70-90% | 90-100% | 0-50% | 50-70% | 70-90% | 90-100% |
| Age |  |  |  |  |  |  |  |  |
| M±SD | 62.05±7.95 | 62.4±7.84 | 62.64±7.72 | 63.55±7.41 | 62.76±7.85 | 61.85±7.91 | 62.01±7.78 | 62.38±7.67 |
| Range | 43.62 - 79 | 43.55 - 78.69 | 43.53 - 78.84 | 43.5 - 78.47 | 43.5 - 78.88 | 43.53 - 79 | 43.69 - 78.76 | 43.84 - 78.41 |
| Sex (% male, N) | 42.24 (18774) | 42.06 (7479) | 43.41 (7718) | 48.94 (4351) | 41.29 (18353) | 44.98 (7998) | 44.71 (7949) | 45.24 (4022) |
| Ethnicity (% white, N) | 96.34 (42668) | 96.98 (17191) | 97.74 (17314) | 98.68 (8750) | 97.7 (43295) | 96.72 (17134) | 96.16 (17032) | 95.56 (8462) |
| Physical activity |  |  |  |  |  |  |  |  |
| M±SD | 27.26±7.91 | 28.19±7.99 | 29.05±8.07 | 30.55±8.46 | 27.88±7.9 | 28.57±8.27 | 28.47±8.24 | 27.84±8.27 |
| Range | 5.12 - 69.28 | 4.94 - 67.94 | 6.46 - 69.41 | 4.83 - 67.35 | 4.83 - 69.21 | 6.46 - 69.28 | 5.88 - 69.2 | 5.12 - 69.41 |
| Employment status (% employed, N) | 63.86 (28201) | 61.76 (10903) | 60.7 (10720) | 56.88 (5020) | 59.67 (26359) | 64.49 (11389) | 65.03 (11474) | 63.74 (5622) |
| Income bracket (M±SD, range 1-5) | 2.86±1.17 | 2.86±1.16 | 2.89±1.16 | 2.9±1.15 | 2.86±1.15 | 2.88±1.17 | 2.9±1.17 | 2.86±1.19 |
| Townsend Deprivation Index |  |  |  |  |  |  |  |  |
| M±SD | -1.58±2.9 | -1.78±2.76 | -1.92±2.69 | -2.25±2.44 | -1.92±2.7 | -1.69±2.82 | -1.61±2.88 | -1.39±2.99 |
| Range | -6.26 - 10.46 | -6.26 - 9.89 | -6.26 - 9.89 | -6.26 - 8.94 | -6.26 - 10.46 | -6.26 - 9.89 | -6.26 - 9.99 | -6.26 - 9.89 |
| Social visits (M±SD, range 1-7) | 5.16±1.12 | 5.19±1.1 | 5.21±1.09 | 5.23±1.09 | 5.18±1.1 | 5.18±1.12 | 5.19±1.11 | 5.17±1.14 |
| Social activities (% >1 weekly, N) | 72.08 (31972) | 72.46 (12857) | 73.58 (13062) | 75.03 (6663) | 72.66 (32249) | 72.89 (12927) | 73.35 (13014) | 71.76 (6364) |
| Smoking |  |  |  |  |  |  |  |  |
| % previous, N | 35.15 (15579) | 36.08 (6396) | 36.87 (6539) | 38.89 (3450) | 34.76 (15408) | 36.06 (6392) | 37.92 (6725) | 38.79 (3439) |
| % current, N | 7.06 (3130) | 7.03 (1247) | 6.58 (1167) | 5.78 (513) | 5.44 (2411) | 7.27 (1288) | 8.18 (1451) | 10.23 (907) |
| Urbanicity (% >10,000 population, N) | 85.44 (36191) | 84.1 (14633) | 83.07 (14482) | 79.29 (6926) | 83.14 (35647) | 83.69 (14380) | 85.51 (14797) | 86.46 (7408) |
| Shift work status (% shift workers, N) | 8.54 (3767) | 8.35 (1473) | 7.32 (1293) | 6.45 (569) | 6.61 (2918) | 8.84 (1560) | 9.82 (1730) | 10.15 (894) |

**Table 2.** Hazard ratios of all-cause and cause-specific mortality for minimally-adjusted Model 1 and fully-adjusted Model 2, alongside percentage and number of deaths, split by light exposure percentiles, and day vs. night.

|  |  | All-cause |  |  |  | Non-cardiomatabolic |  | Cardiomatabolic |  |
| --- | --- | --- | --- | --- | --- | --- | --- | --- | --- |
|  |  | Light intensity percentile | Light intensity range (lx) | % (N) | HR [95%CI] | % (N) | HR [95%CI] | % (N) | HR [95%CI] |
| Model 1 (minimally-adjusted)<br>N = 88,599 | Night | 0-50% (ref.) | <1 | 2.83 (1252) | - | 2.24 (991) | - | 0.58 (255) | - |
|  |  | 50-70% | 1 - 6 | 2.75 (488) | 1.06[0.95-1.17] | 2.17 (384) | 1.05[0.93-1.18] | 0.58 (103) | 1.10[0.87-1.39] |
|  |  | 70-90% | 6 - 48 | 3.05 (541) | **1.16[1.04-1.28]\*\*** | 2.42 (429) | **1.15[1.03-1.29]\*\*** | 0.63 (111) | 1.17[0.94-1.47] |
|  |  | 90-100% | >48 | 3.49 (309) | **1.30[1.15-1.48]\*\*\*** | 2.72 (241) | **1.27[1.10-1.47]\*\*\*** | 0.76 (67) | **1.41[1.07-1.85]\*\*** |
|  | Day | 0-50% (ref.) | <991 | 2.98 (1321) | - | 2.33 (1032) | - | 0.64 (282) | - |
|  |  | 50-70% | 991 - 1750 | 2.88 (510) | 0.92[0.83-1.02] | 2.34 (415) | 0.96[0.85-1.07] | 0.53 (94) | **0.80[0.63-1.01]\*** |
|  |  | 70-90% | 1750 - 3140 | 2.79 (494) | **0.85[0.77-0.95]\*\*** | 2.18 (387) | **0.86[0.76-0.97]\*\*** | 0.60 (106) | 0.86[0.69-1.08] |
|  |  | 90-100% | >3140 | 2.99 (265) | **0.78[0.68-0.89]\*\*\*** | 2.38 (211) | **0.81[0.69-0.94]\*\*** | 0.61 (54) | **0.73[0.54-0.98]\*** |
| Model 2 (fully-adjusted)<br>N = 75,921 | Night | 0-50% (ref.) | <1 | 2.76 (1041) | - | 2.18 (824) | - | 0.56 (212) | - |
|  |  | 50-70% | 1 - 6 | 2.71 (412) | 1.06[0.94-1.19] | 2.09 (318) | 1.01[0.89-1.16] | 0.61 (93) | 1.16[0.91-1.49] |
|  |  | 70-90% | 6 - 48 | 3.07 (472) | **1.15[1.02-1.28]\*** | 2.43 (374) | **1.15[1.02-1.30]\*** | 0.63 (97) | 1.20[0.94-1.54] |
|  |  | 90-100% | >48 | 3.40 (258) | **1.20[1.04-1.38]\*** | 2.65 (201) | **1.15[0.98-1.35]\*** | 0.74 (56) | **1.30[0.95-1.76]\*** |
|  | Day | 0-50% (ref.) | <991 | 2.94 (1104) | - | 2.28 (853) | - | 0.65 (245) | - |
|  |  | 50-70% | 991 - 1750 | 2.79 (426) | 0.94[0.84-1.05] | 2.29 (349) | 1.00[0.88-1.13] | 0.50 (76) | **0.76[0.59-0.99]\*** |
|  |  | 70-90% | 1750 - 3140 | 2.72 (420) | 0.94[0.83-1.05] | 2.13 (329) | 0.95[0.83-1.08] | 0.58 (90) | 0.91[0.71-1.16] |
|  |  | 90-100% | >3140 | 3.02 (233) | 0.98[0.85-1.13] | 2.41 (186) | 1.02[0.86-1.20] | 0.61 (47) | 0.88[0.63-1.21] |

\* $p < .05$ , \*\* $p < .01$ , \*\*\* $p < .001$ . Model 1 was adjusted for age, sex, and ethnicity. Model 2 was adjusted for age, sex, ethnicity, physical activity, socioeconomic status, social visits, smoking status, urbanicity, and shift work.

### Brighter light at night predicted higher risk of all-cause mortality

Exposure to the brightest 10% of night light (00:30 to 06:00) was associated with higher risk of all-cause mortality (minimally-adjusted: HR[95%CI]=1.30[1.15-1.48], p<.001, fully-adjusted: HR=1.20[1.04-1.38], p=.01) when compared to exposure to the dimmest night light (0-50^th^ percentiles; Table 2; Figure 1). Exposure to night light between the 70-90^th^ percentiles was also associated with higher all-cause mortality risk (minimal: HR=1.16[1.04- 1.28]; p=.005, full: HR=1.15[1.02-1.28], p=.02), but exposure between the 50-70^th^ percentiles was not. Light exposure during the day (07:30-20:30) was associated with lower risk of all-cause mortality in the minimal model, for the brightest 10% (HR=0.78[0.68-0.89], p<.001) and between the 70-90^th^ percentiles (HR=0.85[0.77-0.95], p=.003). Day light did not significantly predict mortality risk in fully-adjusted models. Exposure to night light between the 70-90^th^ and 90-100^th^ percentiles were significant predictors of higher mortality risk after additional adjustments for baseline cardiometabolic risk factors, including diabetes diagnosis, vascular diagnoses (heart attack, stroke, angina), hypertension, and high BMI in fully-adjusted models (see Supplementary S7). Mediation models indicated that short sleep duration (<6 h) was a significant predictor of higher all-cause mortality risk (total effect: HR=1.33[1.18-1.49], p<.001), and that light at night was a significant partial mediator of this relationship between short sleep duration and all-cause mortality (direct effect: HR=1.27[1.12-1.44], p<.001; indirect effect: HR=1.04[1.00-1.09], p=.04; see Supplementary S11).

**Figure 1.**
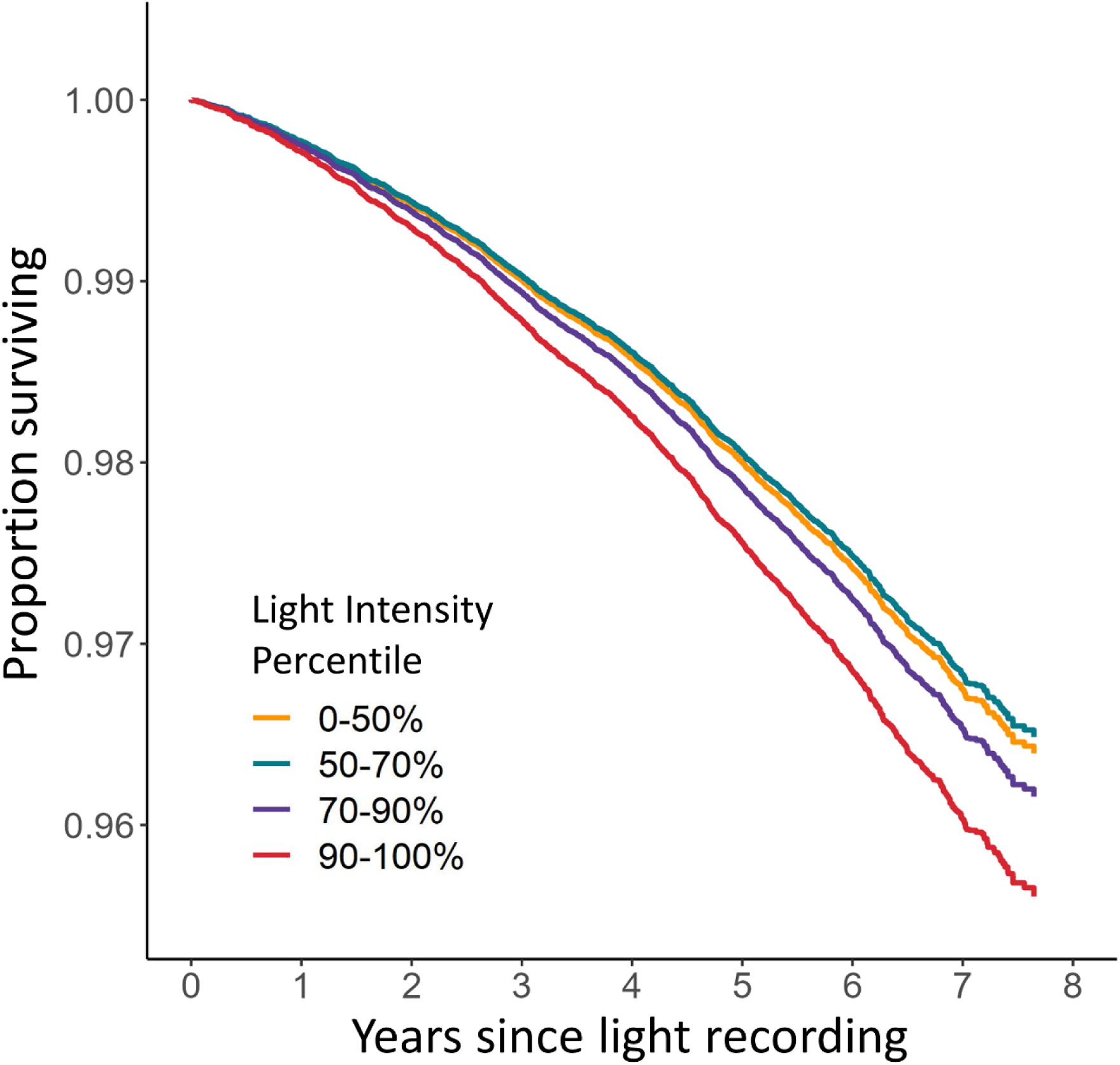
Survival of participants across 7.8 years for 0-50%, 50-70%, 70-90%, and 90-100% light intensity groups, adjusted for age, sex, and ethnicity.

### Light at night and risk of cardiometabolic and non-cardiometabolic mortality

Exposure to the brightest 10% of night light was associated with higher risk of death from cardiometabolic causes when compared to exposure to the dimmest night light (0-50^th^ percentiles; Table 2; Figure 1; minimal: HR[95%CI]=1.41[1.07-1.85], p=.007; full: HR=1.30[0.95-1.76], p=.05). The brightest 10% also had a higher risk of non-cardiometabolic mortality (minimal: HR=1.27[1.10-1.47], p<.001; full: HR=1.15[0.98-1.35], p=.05). Exposure to night light between the 70-90^th^ percentiles was associated with higher non-cardiometabolic mortality risk (minimal: HR=1.15[1.03-1.29], p=.008; full: HR=1.15[1.02-1.30], p=.02) but not higher cardiometabolic mortality risk. Exposure to night light between the 50-70^th^ percentiles was not associated with cardiometabolic or non-cardiometabolic mortality. Day light exposure was associated with lower risk of cardiometabolic mortality (90-100^th^ percentiles: HR=0.73[0.54-0.98], p=.02) and non-cardiometabolic mortality (70-90^th^ percentiles: HR=0.86[0.76-0.97], p=.005; 90-100^th^ percentiles: HR=0.81[0.69-0.94], p=.003) in minimally-adjusted models.

### Time-of-day association of light exposure with all-cause and cardiometabolic mortality

Exposure to the brightest 10% of lighting environments in half-hour intervals between 01:00-06:00 predicted a 23-39% higher risk of all-cause mortality in minimal models, and 18-33% in fully-adjusted (Figure 2), when compared to exposure to the dimmest 0-50^th^ percentiles. The brightest 10% also had a 37-75% higher cardiometabolic mortality risk between 01:00-06:00 in minimal models, and 39-73% between 02:00-06:00 in fully-adjusted. Peak cardiometabolic risk occurred between 02:30-03:30. Non-cardiometabolic risk was numerically lower than cardiometabolic risk in both minimal (14-35% between 00:30-06:00) and fully-adjusted (15-28% between 01:00-06:00) models. Exposure to the brightest 10% of lighting environments between 07:00- 21:00 predicted 12-29% lower risk of all-cause mortality, and exposure between 07:00-21:00 predicted a 14-25% lower risk of non-cardiometabolic mortality, in the minimal but not fully-adjusted models.

**Figure 2.**
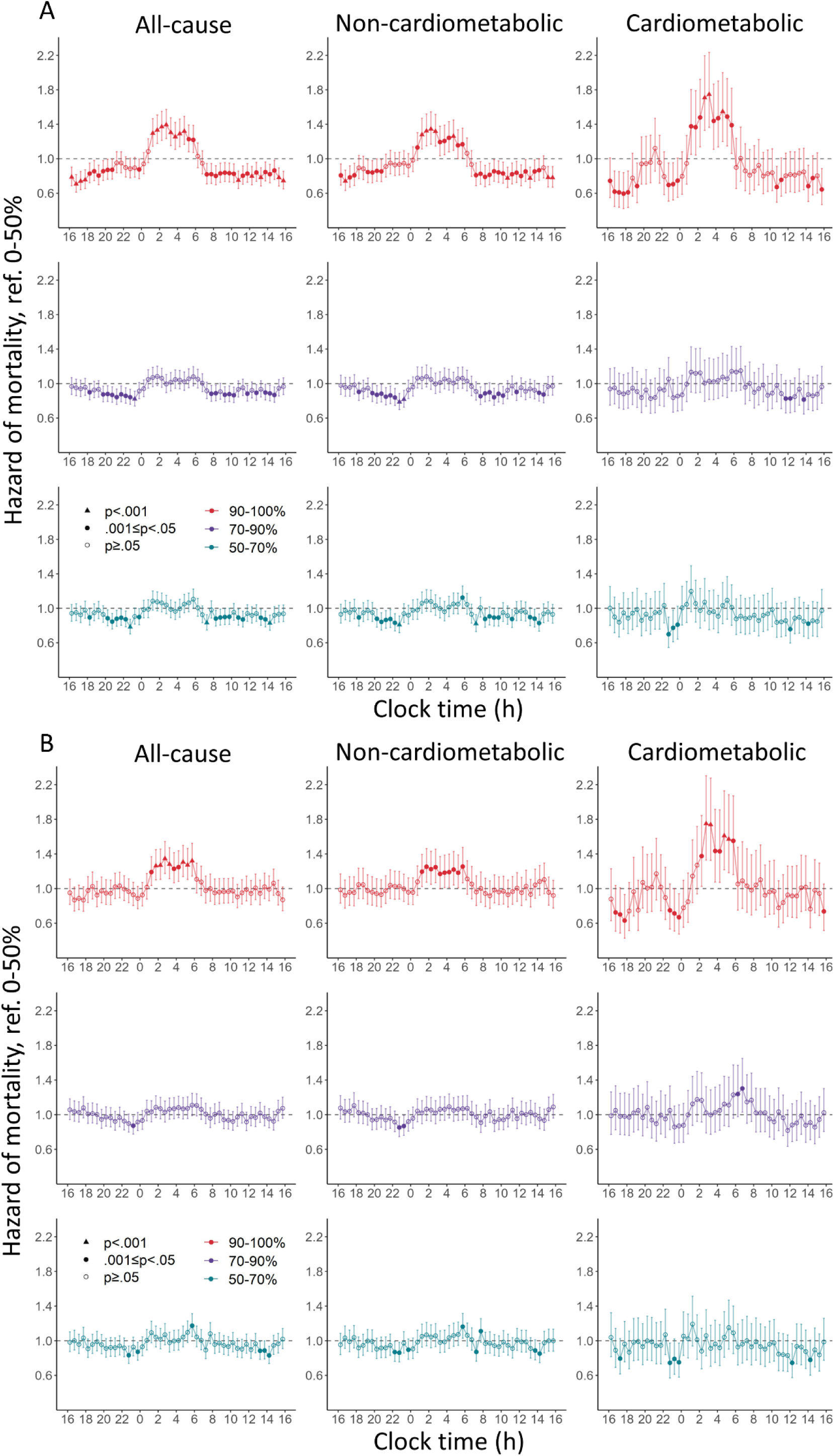
Hazard ratios [95%CI] of all-cause, cardiometabolic, and non-cardiometabolic mortality for light exposures across 24 h, including minimally-adjusted (A) and fully-adjusted (B) models. Separate models were implemented for each half-hour clock time interval, each including 50-70%, 70-90%, and 90-100% light intensity groups referenced against 0-50%. Bonferroni correction for multiple comparisons required p<.001 for statistical significance.

### Lower circadian amplitude and deviated circadian phase predicted higher risk of all-cause and cardiometabolic mortality

Higher minimum circadian amplitude predicted lower risk of all-cause mortality (minimal: HR=0.94[0.91-0.97] per standard deviation, p<.001; full: HR=0.94[0.91-0.98], p=.005), cardiometabolic mortality (minimal: HR=0.90[0.83-0.96], p=.001; full: HR=0.91[0.84-0.98], p=.006), and non-cardiometabolic mortality (minimal: HR=0.96[0.91-1.00], p=.02, full: HR=0.96[0.92-1.01], p=.05). Higher mean circadian amplitude predicted lower risk of all-cause mortality (minimal: HR=0.95[0.92-0.99], p=.01) and cardiometabolic mortality (minimal: HR=0.89[0.82-0.97], p=.004, full: HR=0.91[0.83-1.00], p=.02), but not non-cardiometabolic mortality (see Table 3). Higher maximum circadian amplitude predicted lower risk of all-cause (HR=0.94[0.90-0.98], p=.001) and non-cardiometabolic mortality (HR=0.96[0.92-1.00], p=.02) in minimal models, and cardiometabolic mortality across both model levels (minimal: HR=0.89[0.81-0.96], p=.002; full: HR=0.92[0.84-1.01], p=.04). Early and late circadian phase quintiles predicted higher risks of all-cause (0-20%: HR=1.33[1.17-1.51], p<.001; 20-40%: HR=1.18[1.03-1.34], p=.02; 80-100%: HR=1.19[1.05-1.35], p=.007), cardiometabolic (0-20%: HR=1.48[1.12-1.97], p=.003; 20-40%: HR=1.38[1.03-1.85], p=.02), and non-cardiometabolic mortality (0-20%: HR=1.30[1.13-1.50], p<.001; 20-40%: HR=1.14[0.98-1.32], p=.04; 80-100%: HR=1.18[1.02-1.36], p=.01) compared to the quintile centered at the sample circular mean phase, in minimal models. In fully-adjusted models, early circadian phase predicted higher risks of all-cause (0-20%: HR=1.19[1.04-1.37], p=.01), cardiometabolic (0-20%: HR=1.35[0.99- 1.84], p=.03; 20-40%: HR=1.35[0.99-1.85], p=.03), and non-cardiometabolic (0-20%: HR=1.15[0.98-1.34], p=.04) mortality. Intra-individual variability in circadian phase was not related to mortality. Numbers of all-cause, cardiometabolic, and non-cardiometabolic deaths across the range of each circadian variable are reported in Supplementary S6.

**Table 3.** Hazard ratios of all-cause, non-cardiometabolic, and cardiometabolic mortality for modeled circadian rhythm metrics.

| Model |  | HR [95%CI] |  |  |
| --- | --- | --- | --- | --- |
|  |  | All-cause | Non-cardiometabolic | Cardiometabolic |
| Minimally-adjusted | Mean Amplitude | **0.95[0.92-0.99]\*\*** | 0.97[0.93-1.02] | **0.89[0.82-0.97]\*\*** |
|  | Min. Amplitude | **0.94[0.91-0.97]\*\*\*** | **0.96[0.92-1.00]\*** | **0.90[0.83-0.96]\*\*** |
|  | Max. Amplitude | **0.94[0.90-0.98]\*\*** | **0.96[0.92-1.00]\*** | **0.89[0.81-0.96]\*\*** |
|  | Phase Variability | 0.99[0.95-1.03] | 0.99[0.94-1.04] | 0.98[0.89-1.06] |
|  | Mean Phase |  |  |  |
|  | 0-20% | **1.33[1.17-1.51]\*\*\*** | **1.30[1.13-1.50]\*\*\*** | **1.48[1.12-1.97]\*\*** |
|  | 20-40% | **1.18[1.03-1.34]\*** | **1.14[0.98-1.32]\*** | **1.38[1.03-1.85]\*** |
|  | 60-80% | 1.02[0.89-1.17] | 0.99[0.85-1.15] | 1.22[0.90-1.64] |
|  | 80-100% | **1.19[1.05-1.35]\*\*** | **1.18[1.02-1.36]\*** | 1.26[0.94-1.70] |
| Fully-adjusted | Mean Amplitude | 0.96[0.92-1.01] | 0.99[0.94-1.04] | **0.91[0.83-1.00]\*** |
|  | Min. Amplitude | **0.94[0.91-0.98]\*\*** | **0.96[0.92-1.01]\*** | **0.91[0.84-0.98]\*\*** |
|  | Max. Amplitude | 0.97[0.93-1.02] | 0.99[0.94-1.04] | **0.92[0.84-1.01]\*** |
|  | Phase Variability | 0.99[0.95-1.03] | 0.99[0.95-1.04] | 0.98[0.89-1.07] |
|  | Mean Phase |  |  |  |
|  | 0-20% | **1.19[1.04-1.37]\*** | **1.15[0.98-1.34]\*** | **1.35[0.99-1.84]\*** |
|  | 20-40% | 1.15[0.99-1.33] | 1.12[0.96-1.31] | **1.35[0.99-1.85]\*** |
|  | 60-80% | 0.97[0.83-1.12] | 0.96[0.81-1.13] | 1.08[0.77-1.50] |
|  | 80-100% | 1.06[0.92-1.22] | 1.05[0.89-1.23] | 1.09[0.79-1.51] |

\* p<.05, \*\* p<.01, \*\*\* p<.001. Hazard ratios represent difference in mortality hazard per standard deviation increase in each circadian metric for mean, min., and max. amplitude, and phase variability. Hazard ratios for mean phase represent hazard of each percentile group relative to the 40-60% reference group, centered at the population mean phase (03:50). Phase ranges relative to sample mean for each percentile group were: -12 to -1.16 h (0-20%), -1.16 to -0.40 h (20-40%), 0.40 to 1.16 h (60-80%), and 1.16 to 12 h (80-100%).

## Discussion

Across 14 million hours of light sensor data in ∼89,000 individuals, those with exposure to brighter light at night had a higher risk of all-cause mortality, and higher risk of mortality from cardiometabolic causes. Modeling the impact of light on the circadian system indicated that suppressed circadian amplitude and deviated circadian phase were associated with all-cause and cardiometabolic mortality, consistent with the known biological effects of light exposure at night on the circadian clock.^5^

Exposure to brighter light at night (between 00:30 and 06:00) was associated with higher risk of all-cause mortality. The effects of light were dose-dependent, with higher risk for those with the brightest levels of light at night. Compared to individuals with low night light exposure (the 0-50th percentile), individuals in the 70-90^th^ percentiles of night light exposure had a 15-16% higher risk of all-cause mortality, while individuals in the 90- 100^th^ percentiles of night light exposure had a 20-30% higher risk of all-cause mortality. Associations between light at night and all-cause mortality were robust to multiple levels of adjustment for covarying factors, including age, sex, ethnicity, physical activity, socio-economic advantage, social activity, smoking, urbanicity, shift work, daylight exposure, and baseline cardiometabolic health.

Exposure to brighter light at night was associated with mortality by cardiometabolic and non-cardiometabolic causes. Individuals in the 90-100^th^ percentiles of night light exposure had a 30-41% higher risk of cardiometabolic mortality, compared to those with low night light exposure (the 0-50^th^ percentile). In comparison, for non-cardiometabolic mortality, individuals in the 90-100^th^ percentiles of night light exposure had a 15-27% higher risk. These findings are consistent with the role of light exposure at night in the development of metabolic syndrome and obesity,^32–36^ and with the role of circadian disruption in the development of cardiometabolic diseases including myocardial infarction, stroke, hypertension, and diabetes.^19–22^ Analysis of mortality risk for light exposure across half-hour intervals showed a peak cardiometabolic risk between 02:30-03:00 (73% greater risk for brightest 10% vs. bottom 50%). This is consistent with evidence that circadian rhythms are most disrupted by light exposure across a short interval in the middle of the biological night,^5,37^ and also consistent with our modeling results that link lower circadian amplitude to higher cardiometabolic mortality risk.

Light synchronizes the timing of the brain’s central circadian pacemaker to the 24 h light/dark cycle, but light exposure during the night also causes suppression of circadian amplitude and shifted circadian phase.^4,5,38^ Using a validated computational model representing the dynamic response of the central circadian clock to light, we found that disrupted circadian rhythms predicted higher mortality risk. Each standard deviation reduction in circadian amplitude was associated with a 5-6% higher all-cause mortality risk and a 9-11% higher cardiometabolic mortality risk. Individuals whose circadian phase minima occurred more than one hour before the group average had a 19-33% higher risk of all-cause mortality, and a 35-48% higher risk of cardiometabolic mortality. These findings support the notion that circadian disruption is the mechanism linking light exposure at night with higher mortality risk. This link could be explained by the role of circadian disruption in the initiation and progression of disease,^1^ or by the disruption of circadian regulation in gene expression that correlates with premature mortality.^39^

This study investigated the relationship between individual-level light exposure and mortality risk in a large, well-characterized cohort, using personal light sensors. Previous large-scale studies have assessed satellite-derived outdoor light exposure, finding associations with risk of all-cause mortality, and risk of coronary heart disease.^15,16^ However, satellite data captures the outdoor environment only, and may not be an ideal proxy for an individual’s light exposure pattern, including indoor light levels.^40^ Our analyses use data from personal sensors and therefore capture a range of lighting environments specific to each individual, which is especially important at night, when individuals are most at risk of exposure to light that disrupts their circadian rhythms.^4,5^ Personal sensor data also allows for inclusion of individual-level day light exposure in mortality risk models, an important control given day light can alter the sensitivity of the circadian system to light at night.^41–43^ Furthermore, individual-level data allowed us to model the effect of light exposure on each individual’s circadian system, an approach that incorporates information about their light exposure history. This modeling approach provides mechanistic insight into the links between light at night, circadian rhythms, and mortality, and expands upon research that derived circadian metrics from accelerometer data only.^13,14^

There are several limitations to this study. Firstly, only one week of light exposure was available for each participant. Light patterns, however, were stable across up to four repeated-measures collections in ∼3,000 participants, indicating that one week of data was a reasonable proxy for an individual’s typical light patterns. Secondly, light/activity recordings did not occur simultaneously with collection of several covariates that are subject to change over time. Thirdly, the computational model was developed using studies of healthy younger adults,^5,44^ and does not account for individual differences in physiology, including possible age-related changes.^45^

Finally, since this is a correlational study, is it possible that brighter light at night and premature mortality are caused by unmeasured factors. However, evidence supports a causal pathway from night light exposure to circadian disruption and premature mortality, particularly for cardiometabolic mortality.

These findings demonstrate the importance of maintaining a dark environment across the late night and early morning hours, when the central circadian pacemaker is most sensitive to light. Protection of night lighting environments may be especially important in those at risk for both circadian disruption and mortality, for example in intensive care or aged care settings.^46,47^ Across the general population, avoiding light at night may lead to reduction in disease burden, especially cardiometabolic diseases, and may enhance longevity.

## Supporting information

STROBE Checklist

## Data sharing statement

The data underlying this work are available at the UK Biobank website, upon application: https://www.ukbiobank.ac.uk/enable-your-research/apply-for-access. Scripts for data handling and analysis will be made available upon request by Daniel P. Windred.

## Acknowledgement

This research has been conducted using data from UK Biobank, a major biomedical database (Project ID: 6818). We thank the UK Biobank study participants, and the UK Biobank team for their work in developing and maintaining this resource.

DPW, SWC, and AJKP conceived the study, and drafted the first manuscript. DPW coordinated the research, under supervision from AJKP and SWC, and in partnership with AB. DPW was responsible for data handling, cleaning, and analysis, with contributions from AB. All authors contributed to the interpretation and discussion of findings. All authors revised and approved the final version of the manuscript. DPW had full access to all the data in the study and takes responsibility for the integrity of the data and the accuracy of the data analysis.

AJKP and SWC received research funding from Versalux and Delos, and are co-founders and co-directors of Circadian Health Innovations PTY LTD. SWC has also consulted for Dyson, and received research funding from Beacon Lighting. PO co-founded Axivity Ltd, and was a Director until 2015. DPW: none; ACB: none; RS: none; JML: none; MKR: none. This work was supported by the NIHR Manchester Biomedical Research Centre.

