## Supplementary Material for "Light at night and modeled circadian disruption predict higher risk of mortality: A prospective study in >88,000 participants"

**Affiliation / Institution:**

[1]Turner Institute for Brain and Mental Health, School of Psychological Sciences, Faculty of Medicine, Nursing and Health Sciences, Monash University, Melbourne, VIC, Australia

[2]Center for Genomic Medicine, Massachusetts General Hospital, Boston, MA, USA

[3]Department of Anesthesia, Critical Care and Pain Medicine, Massachusetts General Hospital and Harvard Medical School, Boston, MA, USA

[4]Division of Sleep and Circadian Disorders, Brigham and Women's Hospital, Boston, MA, USA

[5]Action Lab, Department of Human-Centred Computing, Faculty of Information Technology, Monash University, Melbourne, VIC, Australia

[6]Centre for Biological Timing, Division of Endocrinology, Diabetes & Gastroenterology, School of Medical Sciences, Faculty of Biology, Medicine and Health, Manchester Academic Health Science Centre, University of Manchester, Manchester, UK

[7]Diabetes, Endocrinology and Metabolism Centre, NIHR Manchester Biomedical Research Centre, Manchester University NHS Foundation Trust, Manchester, UK.

*Authors contributed equally to this manuscript.

**Corresponding Author:** Sean W. Cain

**Table S1.** Links to UK Biobank protocol documents

| Document | Link |
|---|---|
| Invitation to participate | https://biobank.ctsu.ox.ac.uk/crystal/refer.cgi?id=100253 |
| Death registry linkage | https://biobank.ctsu.ox.ac.uk/crystal/refer.cgi?id=115559 |
| Light / accelerometer: Participant instructions | https://biobank.ndph.ox.ac.uk/showcase/refer.cgi?id=141141 |
| Light / accelerometer: Collection and processing | https://biobank.ndph.ox.ac.uk/showcase/refer.cgi?id=131600 |
| Assessment centre: Consent | https://biobank.ndph.ox.ac.uk/showcase/refer.cgi?id=100230 |
| Assessment centre: Reception | https://biobank.ndph.ox.ac.uk/showcase/ukb/docs/Reception.pdf |
| Assessment centre: Blood pressure | https://biobank.ndph.ox.ac.uk/showcase/refer.cgi?id=100225 |
| Assessment centre: Blood biochemistry | https://biobank.ndph.ox.ac.uk/showcase/refer.cgi?id=5636<br>https://biobank.ndph.ox.ac.uk/showcase/refer.cgi?id=1227 |

**Table S2.** UK Biobank variables

| Variable | Collection method | UKB ID | Description | Link |
|---|---|---|---|---|
| Age | Registry | 21003 | Participant age (years) at assessment centre visit. Data obtained from NHS Primary Care Trust registries. Confirmed with participants at assessment centre, and amended if required. | https://biobank.ndph.ox.ac.uk/showcase/field.cgi?id=21003 |
| Sex | Registry | 31 | Obtained from NHS Primary Care Trust registries. Confirmed with participants at assessment centre, and amended if required. | https://biobank.ndph.ox.ac.uk/showcase/field.cgi?id=31 |
| Ethnic background | Assessment centre visit: Questionnaire | 21000 | Ethnic group (white, mixed, Asian/Asian British, black/black British, Chinese, other, PNTA) and ethnic background (sub-categories within each ethnic group). | https://biobank.ndph.ox.ac.uk/showcase/field.cgi?id=21000 |
| Current employment status | Assessment centre visit: Questionnaire | 6142 | Paid employment, unemployed, retired, home/family caretaker, unable to work, volunteer, student, other | https://biobank.ndph.ox.ac.uk/showcase/field.cgi?id=6142 |
| Average total household income before tax | Assessment centre visit: Questionnaire | 738 | Income brackets: <£18,000, £18,000-£29,900, £30,000-£51,900, £52,000-£100k,000, >£100,000, DNK, PNTA | https://biobank.ndph.ox.ac.uk/showcase/field.cgi?id=738 |
| Townsend deprivation index | Registry | 189 | Scores represent average home ownership, car ownership, household overcrowding, and employment rate across each participant's postcode. Calculated using national census data at time of recruitment. | https://biobank.ndph.ox.ac.uk/showcase/field.cgi?id=189 |
| Leisure / social activities | Assessment centre visit: Questionnaire | 6160 | Participation in one or more weekly social activities, including: sports club /gym, pub or social club, adult education, other group activity, none, PNTA | https://biobank.ndph.ox.ac.uk/showcase/field.cgi?id=6160 |
| Frequency of friend/family visits | Assessment centre visit: Questionnaire | 1031 | Frequency of visits from family/friends: Almost daily, 2-4 times a week, once a week, once a month, once every few months, never/almost never, no friends/family outside household, DNK, PNTA | https://biobank.ndph.ox.ac.uk/showcase/field.cgi?id=1031 |
| Smoking status | Assessment centre visit: Questionnaire | 20116 | Smoking status / history: never, previous, current, PNTA | https://biobank.ndph.ox.ac.uk/showcase/field.cgi?id=20116 |
| Urbanicity | Registry | 20118 | 'Urban' (population ≥ 10,000) and 'non-urban' (population < 10,000), defined according to population density of participant's postcode, derived from the UK Office for National Statistics. | https://biobank.ndph.ox.ac.uk/showcase/field.cgi?id=20118 |
| Shift work | Assessment centre visit: Questionnaire | 826 | 'Does your work involve shift work?' Answers: never, sometimes, usually, always, DNK, PNTA | https://biobank.ndph.ox.ac.uk/showcase/field.cgi?id=826 |
| Diabetes status | Assessment centre visit: Questionnaire | 2443 | 'Has a doctor ever told you that you have diabetes?' Answers: Yes, no, DNK, PNTA | https://biobank.ndph.ox.ac.uk/showcase/field.cgi?id=2443 |

| | | | | |
|---|---|---|---|---|
| Vascular condition diagnosed by a doctor | Assessment centre visit: Questionnaire | 6150 | ‘Has a doctor ever told you that you have had any of the following conditions?’ Answers: Heart attack, angina, stroke, high blood pressure, none, PNTA | https://biobank.ndph.ox.ac.uk/showcase/field.cgi?id=6150 |
| Body mass index | Assessment centre visit: Physical | 21001 | Weight (kg) / height (m)^2 | https://biobank.ndph.ox.ac.uk/showcase/field.cgi?id=21001 |
| High density lipoprotein | Assessment centre visit: Physical | 30760 | Blood biochemistry assay | https://biobank.ndph.ox.ac.uk/showcase/field.cgi?id=30760 |
| Low density lipoprotein | Assessment centre visit: Physical | 30780 | Blood biochemistry assay | https://biobank.ndph.ox.ac.uk/showcase/field.cgi?id=30780 |
| Triglycerides | Assessment centre visit: Physical | 30870 | Blood biochemistry assay | https://biobank.ndph.ox.ac.uk/showcase/field.cgi?id=30870 |
| Systolic blood pressure | Assessment centre visit: Physical | 4080 | Two readings, recorded at the start and end of assessment centre visit | https://biobank.ndph.ox.ac.uk/showcase/field.cgi?id=4080 |
| Diastolic blood pressure | Assessment centre visit: Physical | 4079 | Two readings, recorded at the start and end of assessment centre visit | https://biobank.ndph.ox.ac.uk/showcase/field.cgi?id=4079 |
| Physical activity | Accelerometer | 90012 | Axivity AX3 device average acceleration across one week data collection | https://biobank.ndph.ox.ac.uk/showcase/field.cgi?id=90012 |

DNK = ‘do not know’; PNTA = ‘prefer not to answer’

**Table S3.** Covariates included in statistical models, as derived from UK Biobank variables

| Covariate | UKB ID(s) | Description |
|---|---|---|
| Age | 21003 | Strata, split by quartiles: Q1 [43.3-56.0], Q2 [56.0-63.4], Q3 [63.4-68.7], Q4 [68.7-82.0] (years at light/actigraphy recording) |
| Sex | 31 | Binary, male = 1, female = 0 |
| Ethnicity | 21000 | Binary, white ethnic group = 1, other ethnic group = 0 |
| Physical activity | 90012 | Strata, split by quartiles: Q1 [0-21.9], Q2 [21.9-26.8], Q3 [26.8-32.4], Q4 [32.4-69.4] (milli-gravity) |
| Employment status | 6142 | Binary, employed = 1, other categories = 0 |
| Income | 738 | Ordered categories, <£18k = 1, £18k-£29.9k = 2, £30k-£51.9k = 3, £52k-£100k = 4, and >£100k = 5. |
| Deprivation | 189 | Continuous, included as recorded |
| Social activities | 6160 | Binary, >0 weekly activities = 1, 0 weekly activities = 0 |
| Social visits | 1031 | Ordered categories: 'no friends / family' = 1 'never or almost never' = 2, 'every few months' = 3, 'monthly' = 4, 'weekly' = 5, '2-4 times a week' = 6, and 'almost daily' = 7 |
| Smoking status | 20116 | Categorical, referent category = 'never' |
| Urbanicity | 20118 | Binary, urban = 1, rural = 0 |
| Shift work | 826 | Binary, 'Always' = 1, all other categories = 0 |
| Diabetes status | 2443 | Binary strata, 'Yes' = 1, 'No' = 0 |
| Vascular diagnosis | 6150 | Binary strata, 'Heart attack', and/or 'Stroke', and/or 'Angina', 'None' = 0 |
| BMI | 21001 | Binary strata, BMI > 30 = 1, BMI ≤ 30 = 0 |
| Cholesterol ratio | 30760, 30780, 30870 | Binary strata, 'high' = cholesterol ratio > 3.75 for males or > 3.00 for females. Calculated as cholesterol ratio = (HDL + LDL + 0.2*triglycerides)/HDL |
| Hypertension | 4080, 4079 | Binary strata, 'high' = systolic > 140 or diastolic > 90, where systolic and diastolic were included as the average of two readings (mmHg) |

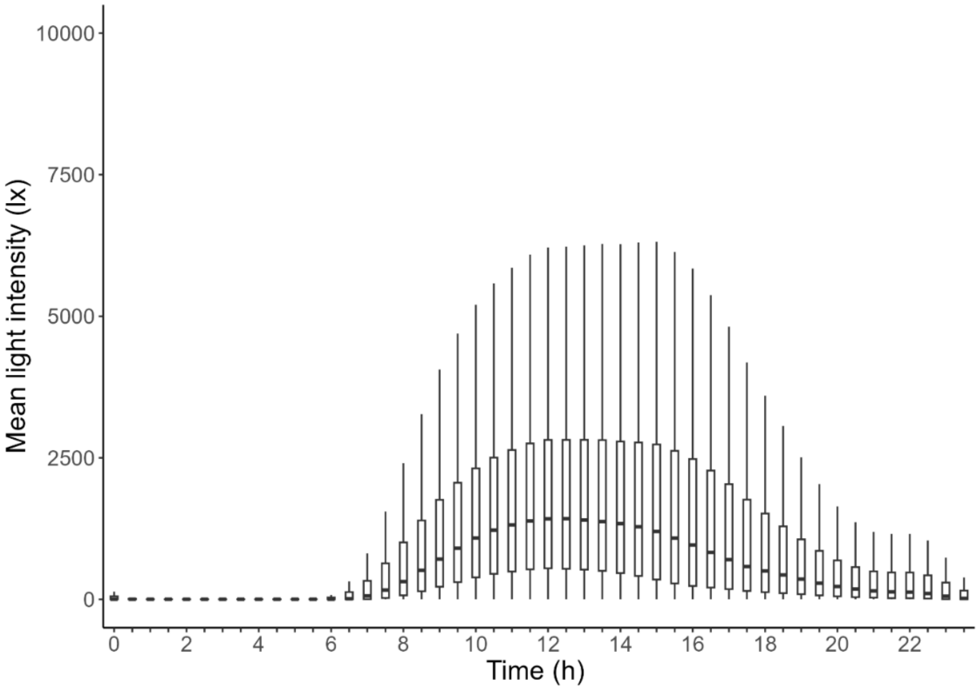

**Figure S4.** Approximated mean light intensity by half-hour clock time intervals

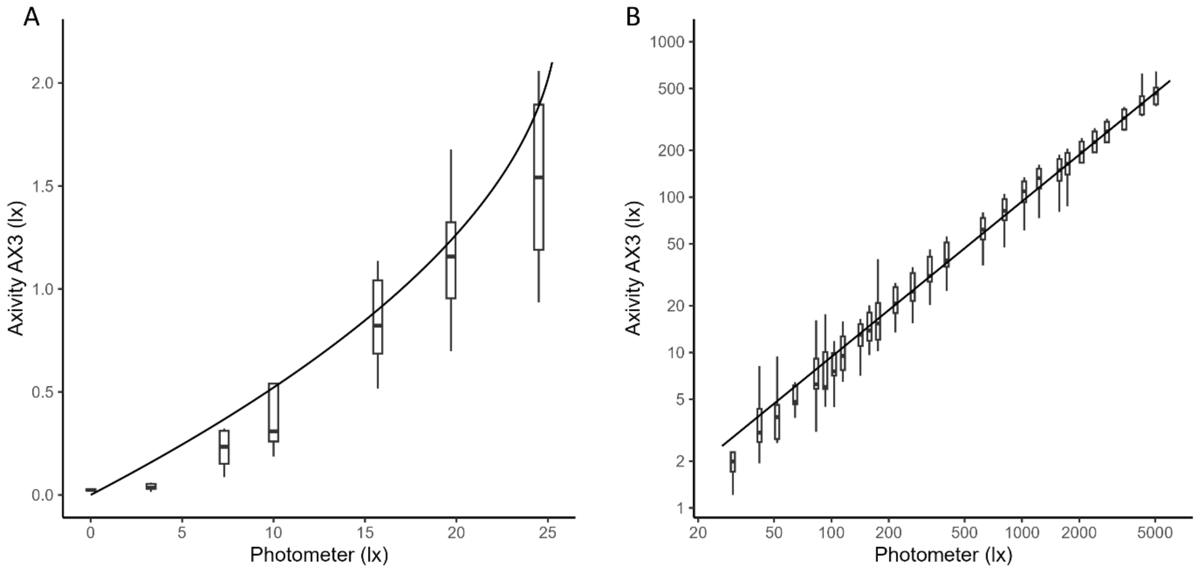

**Figure S5.** AX3 validation: light intensity for device output vs. reference photometer. Test data were split into (A) 0-25lx photometer range (non-linear device response), and (B) 25-5500lx photometer range (linear device response)

**Table S6.** Percentage and number of deaths in quintiles of each modeled circadian variable

| | | | Deaths % (N) in circadian variable quintiles | | | | |
|---|---|---|---|---|---|---|---|
| | **Circadian variable** | | Q1 | Q2 | Q3 | Q4 | Q5 |
| Model 1 | Mean Amplitude | All-cause | 3.16(541) | 2.79(478) | 2.80(480) | 2.82(484) | 2.82(505) |
| (N = 88,599) | | Non-cardiometabolic | 0.78(134) | 0.50(86) | 0.53(91) | 0.63(108) | 0.63(96) |
| | | Cardiometabolic | 2.36(405) | 2.25(386) | 2.27(389) | 2.19(375) | 2.19(409) |
| | Min· Amplitude | All-cause | 3.13(537) | 2.76(473) | 2.61(447) | 2.82(484) | 2.82(547) |
| | | Non-cardiometabolic | 0.69(119) | 0.60(102) | 0.53(91) | 0.51(88) | 0.51(115) |
| | | Cardiometabolic | 2.43(417) | 2.14(367) | 2.05(352) | 2.31(396) | 2.31(432) |
| | Max· Amplitude | All-cause | 3.12(534) | 2.92(501) | 2.77(474) | 3.05(522) | 3.05(457) |
| | | Non-cardiometabolic | 0.73(125) | 0.57(97) | 0.59(101) | 0.61(104) | 0.61(88) |
| | | Cardiometabolic | 2.37(406) | 2.35(402) | 2.16(370) | 2.43(417) | 2.43(369) |
| | Average Phase | All-cause | 3.50(600) | 2.84(487) | 2.43(417) | 2.58(443) | 2.58(541) |
| | | Non-cardiometabolic | 0.75(128) | 0.62(106) | 0.45(77) | 0.57(97) | 0.57(107) |
| | | Cardiometabolic | 2.75(471) | 2.21(379) | 1.96(336) | 2.02(346) | 2.02(432) |
| | Phase Variability | All-cause | 3.07(526) | 3.14(538) | 2.91(499) | 2.66(455) | 2.66(470) |
| | | Non-cardiometabolic | 0.62(107) | 0.67(114) | 0.61(104) | 0.50(85) | 0.50(105) |
| | | Cardiometabolic | 2.43(416) | 2.46(422) | 2.31(395) | 2.16(370) | 2.16(361) |
| Model 3 | Mean Amplitude | All-cause | 3.15(447) | 2.78(416) | 2.84(425) | 2.72(408) | 2.84(420) |
| (N = 75,921) | | Non-cardiometabolic | 0.80(113) | 0.54(81) | 0.53(80) | 0.58(87) | 0.55(81) |
| | | Cardiometabolic | 2.34(332) | 2.20(330) | 2.30(345) | 2.14(320) | 2.29(339) |
| | Min· Amplitude | All-cause | 3.11(462) | 2.73(401) | 2.72(401) | 2.70(402) | 3.05(450) |
| | | Non-cardiometabolic | 0.71(105) | 0.59(87) | 0.56(82) | 0.49(73) | 0.64(95) |
| | | Cardiometabolic | 2.40(356) | 2.11(310) | 2.14(316) | 2.21(329) | 2.41(355) |
| | Max· Amplitude | All-cause | 3.09(434) | 2.90(434) | 2.77(414) | 2.98(447) | 2.58(387) |
| | | Non-cardiometabolic | 0.74(104) | 0.59(88) | 0.58(87) | 0.58(87) | 0.51(76) |
| | | Cardiometabolic | 2.32(327) | 2.31(345) | 2.17(324) | 2.39(359) | 2.08(311) |
| | Average Phase | All-cause | 3.46(508) | 2.91(434) | 2.40(358) | 2.48(369) | 3.09(447) |
| | | Non-cardiometabolic | 0.78(115) | 0.64(96) | 0.45(67) | 0.51(76) | 0.61(88) |
| | | Cardiometabolic | 2.67(392) | 2.25(336) | 1.93(288) | 1.97(293) | 2.46(357) |
| | Phase Variability | All-cause | 3.01(434) | 3.10(455) | 2.85(422) | 2.72(408) | 2.63(397) |
| | | Non-cardiometabolic | 0.58(84) | 0.70(102) | 0.62(91) | 0.53(79) | 0.57(86) |
| | | Cardiometabolic | 2.41(347) | 2.39(351) | 2.24(331) | 2.20(329) | 2.04(308) |

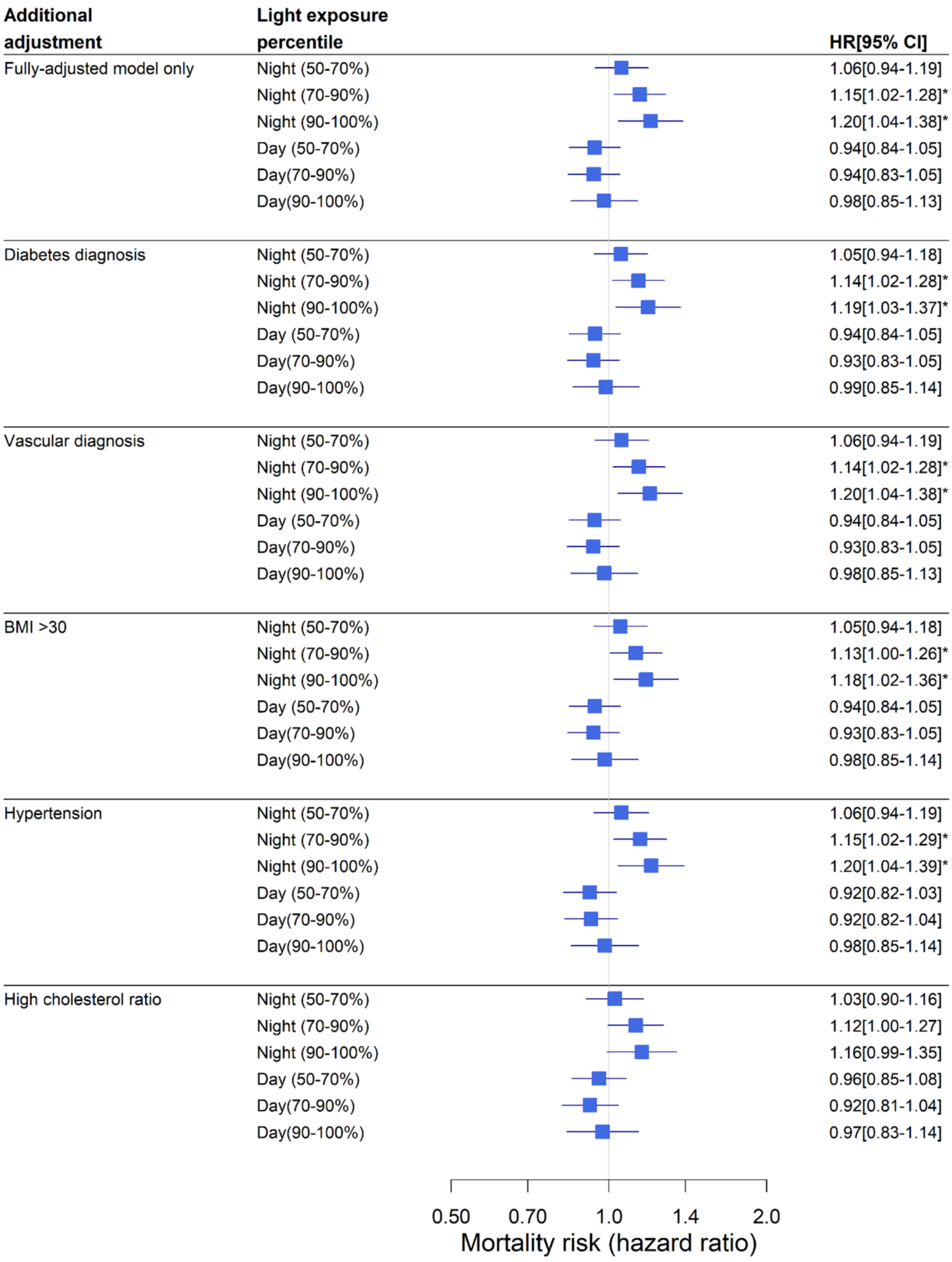

**Figure S7.** Light at night and all-cause mortality, adjusted for baseline cardiometabolic health

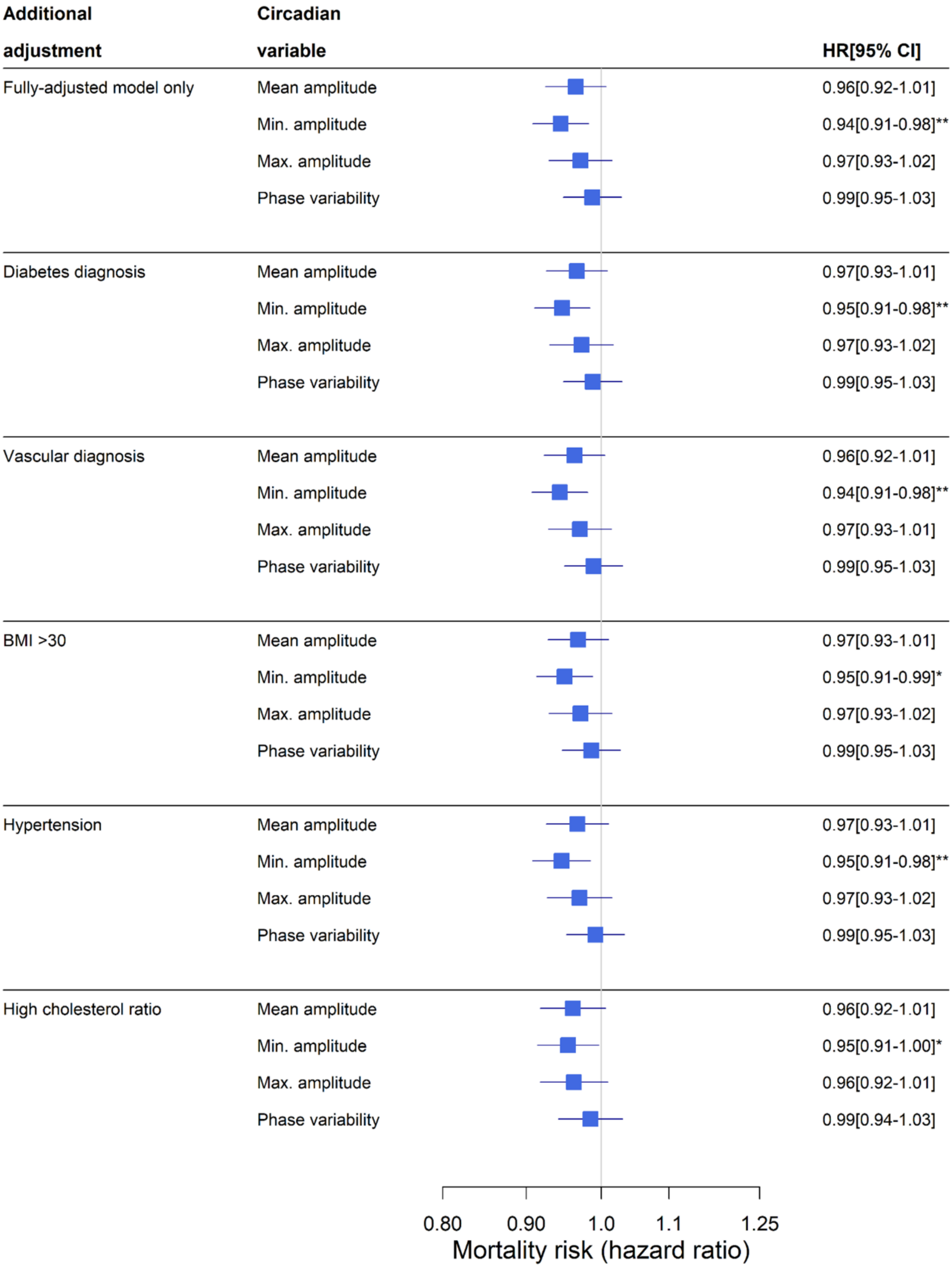

**Figure S8.** Modeled circadian variables and all-cause mortality, adjusted for baseline cardiometabolic health

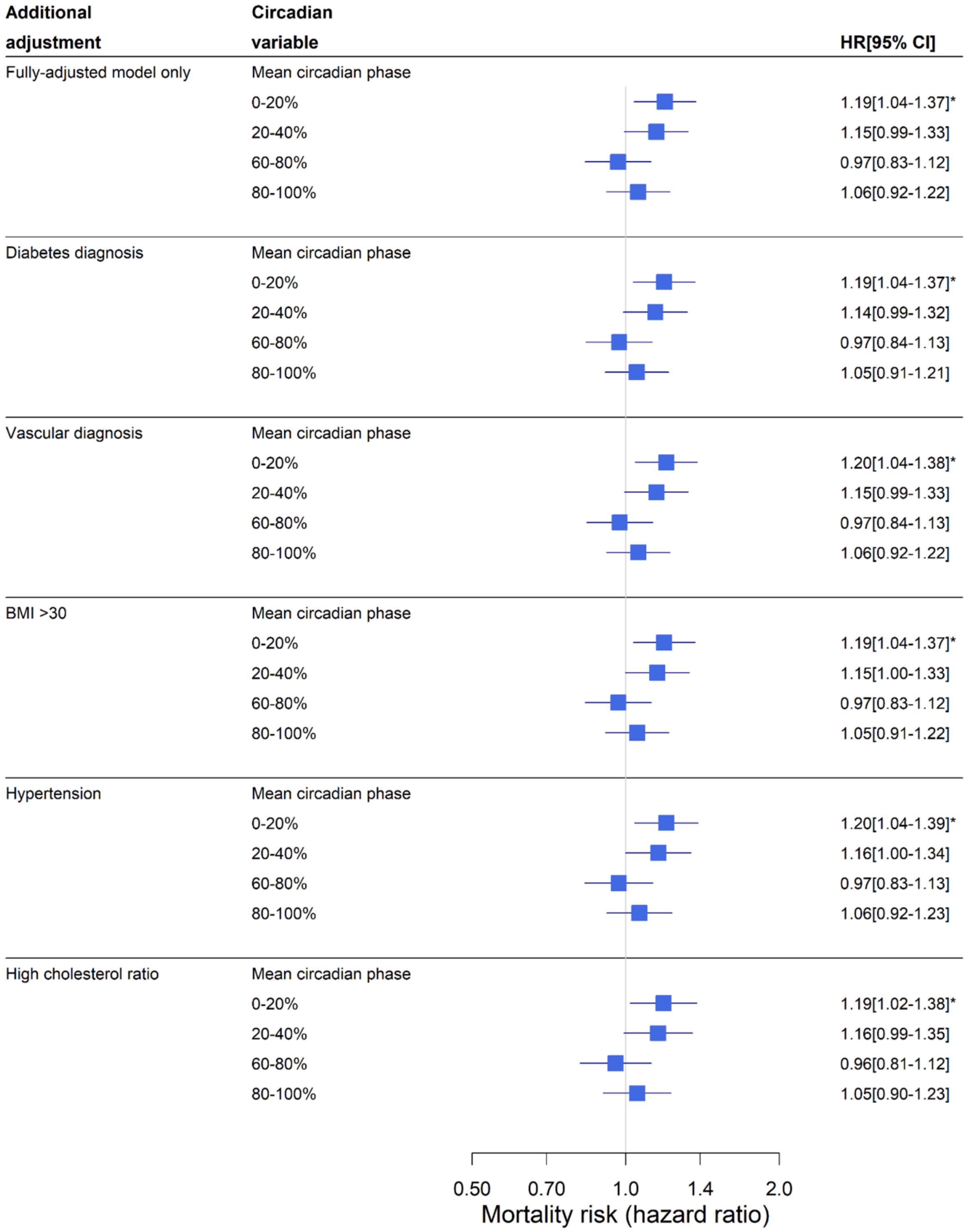

**Figure S9.** Modeled circadian phase and all-cause mortality, adjusted for baseline cardiometabolic health

### S10. Supplementary Methods

#### Protocol

The initial cohort of approximately 502,000 participants underwent assessment at one of twenty-two assessment centres across the UK, located to capture the range of socio-economic, ethnic, and urban-rural distributions across the UK population, between 2006-2010. Invitations to participate in activity/light tracking were sent to 236,519 individuals from the initial cohort via post, and 103,669 agreed to participate, between 2013-2016. Documentation relating to consent and recruitment are available on the UK Biobank website, and relevant links are provided in Table S10.

#### Cox Proportional Hazards Implementation

Cox proportional hazards models were implemented using the 'survival' package in R (version 4.1.0), and competing risks models using the 'crrSC' package. Survival curves were adjusted using the 'survminer' package implementing a 'marginal' approach, which balances each subgroup across all covariates. Contrasts were set using the default 'contr.treatment' setting in R for all categorical variables. This method defines a referent category to which all other categories are compared (e.g., 0-50% light exposure category defined as referent, 50-70%, 70-90%, 90-100% exposure categories as comparison groups).

An example of implemented model syntax for cox proportional hazards models is as follows:

*coxph(Surv(years, death) ~ light_night + light_day + age + sex + ethnicity, data=data)*

where *years* represents time between light recording and either death or censoring, and *death* is a binary marker of participant survival.

An example of model syntax for proportional sub-hazards models is as follows:

*crrs(ftime = years, fstatus = death_type, cov1 = covs, failcode = n, strata = strata_covs, ctype = 1)*

where *years* represents time between light recording and either death or censoring, *death_type* is a numerical representation of cause of death (0 = living, 1 = cardiometabolic, 2 = non-cardiometabolic), *covs* is data structure containing required covariates (e.g., age, sex, ethnicity), *n* represents the cause of death defined as the competing hazard in each model (e.g., n = 1 to test the competing hazard of cardiometabolic mortality), and *strata_covs* represents covariates included as 'strata' variables.

The assumption of proportional hazards was assessed by inspecting the plotted temporal stability of regression coefficients associated with each covariate, using the 'cox.zph' function. Multi-collinearity was assessed by calculating the Variance Inflation Factor (VIF) for all model covariates using the 'ols_vif_tol' function. Age, physical activity, diabetes status, vascular diagnoses, BMI, hypertension, and cholesterol ratio were included in all models as 'strata' variables, accounting for group-wise differences in baseline mortality risk.

#### Light Data Cleaning

Raw data from AX3 devices were initially downsampled to 1Hz, converted to device lux based on the device manual, where device lux = 10^(device output current/341), and averaged into 10-second mean epochs. After cleaning of light data according to non-wear, light data were smoothed by taking the median of all epochs within 10-minute rolling windows, stepped in 2-minute increments, to increase speed of data processing.

#### Light Sensor Testing

We attained a sample of Axivity AX3 devices worn by UK Biobank participants and assessed them against reference lighting conditions. Devices were exposed to intensities between 0-5500 lx, from a point source oriented along their z-axis. Reference light intensity was measured using a Tektronix J17 photometer (Luma Color) oriented vertically towards the point source. Devices captured light down to a minimum between 3-7 lux. The AX3 light sensor exhibited a linear relationship with the reference light source between 25-5,050 lux, and a non linear relationship between 0-25 lux. A piece-wise conversion function was therefore extracted, as follows:

$$I_{approx} = \begin{cases} 21.55 I_{device} - 4.54 {I_{device}}^2, & I_{device} \leq 2.4 \\ 10.67 I_{device}, & I_{device} > 2.4 \end{cases}$$

where $I_{device}$ represents device output light intensity (lux), and $I_{approx}$ represents approximate light intensity (lux) according to the Tektronix J17 photometer.

**Night Light and Day Light Factors**

Night light (00:30-06:00) and day light (07:30-20:30) groupings were extracted using factor analysis, based on factor loading ≥0·5 (varimax rotation, cumulative proportion variance explained = 0·56). Day and night light factors were internally consistent (Cronbach's α=·98 and α=·93, respectively), and exhibited a weak positive correlation ($r_s$=0·10, p<·0001).

**Covariates**

Variables known to influence mortality and light exposure were chosen as model covariates (Table S3). Age, sex, and ethnicity were included in minimally adjusted Model 1, as trait characteristics that may only exhibit a unidirectional causal relationship with both mortality and light exposure. Additional covariates (physical activity, employment, income, deprivation, social visits, social activities, smoking, urbanicity, shift work) were selected due to their possible confounding effect on the mortality/light exposure relationship. These covariates were also potential mediators of the mortality/light exposure relationship, in contrast with age, sex, and ethnicity which were not. All covariates except physical activity (UKB ID: 90012) were collected between 2006-2010 via touch-screen questionnaire and physical collection within a single assessment centre visit, and were therefore subject to change between time of collection and light/activity recoding. Shift work was included as an interaction variable in Model 2.

Sleep was derived using GGIR[1,2], a validated open-source R package for estimating sleep-wake state from accelerometer data. This package was implemented as reported previously, where probable miscalculations of sleep times were identified and associated nights of data were removed[3]. Sleep duration (M ± SD = 6·79 ± 1·05 h) was calculated as the total duration of sustained inactivity between nightly 'sleeponset' and 'wakeup' times estimated by GGIR, where sustained inactivity represented <5° of deviation of the accelerometer from its z-axis for >5 min. The mean of sleep duration was calculated using all available nights in each participant, after excluding all participants with <3 nights of sleep data output by GGIR.

**Circadian Rhythm Modeling**

Light data was input to a dynamic stimulus processer that represents transduction and transmission of light from the environment to the central pacemaker, via photoreceptors and the retino-hypothalamic tract (Process L). Output from Process L was input to a limit cycle oscillator, which represents the central pacemaker (Process P).

Process L was modeled by:

$$\dot{n} = C(\alpha(1-n) - \beta n)$$

$$\alpha = \alpha_0 \left(\frac{I}{I_0}\right)^p$$

where $n$ represents the fraction of saturated photoreceptor elements, and $I$ represents light intensity.

The central pacemaker (Process P) was modeled by:

$$\dot{x} = \frac{\pi}{12}\left(x_c + \hat{B}(1-0.4x)(1-0.4x_c)\right)$$

$$\dot{x}_c = \frac{\pi}{12}\left(\mu\left(x_c - \frac{4x_c^3}{3}\right) - x\left(\left(\frac{24}{0.99729\tau_x}\right)^2 + k\hat{B}(1-0.4x)(1-0.4x_c)\right)\right)$$

$$\hat{B} = G(1-n)\alpha$$

where $x$ and $x_c$ represent the state of the central circadian pacemaker, and $\hat{B}$ represents the drive on the pacemaker of transmission of light information from Process L.

Light intensity $(I)$, photoreceptor state $(n)$, and circadian pacemaker state $(x, x_c)$ are all functions of time, defined at all epochs of each participant's light data recording.

Fixed parameters:

$C = 60$

$\beta = 0.013\ min^{-1}$

$\alpha_0 = 0.16\ min^{-1}$

$I_0 = 9500$

$p = 0.6$

$\mu = 0.23$

$G = 19.875$

$\tau_x = 24.2$

$k = 0.55$

Amplitude was calculated by:

$$A = \sqrt{x^2 + x_c^2}$$

for pairs of $x$ and $x_c$ at all epochs. Mean, minimum, and maximum amplitude were calculated from amplitude across all epochs.

Phase was calculated as clock time at the minimum value of $x$ in each 24h interval, plus a reference value of 0.8h. Phase was extracted for each available day of data, and mean and standard deviation were calculated across all days for each participant.

This modeling approach assumed all input epochs were consecutive and of equal length, but light data contained missing epochs due to device non-wear. We imputed any length of missing light data ≤120 min using Kalman imputation with the 'imputeTS' package in R. After this step, we extracted the longest continuous interval of light data for each participant containing whole days only, and excluded all participants with <3 continuous days of light data (95,068 remaining after exclusion). Participants without valid daily light profiles were also excluded, as described in the main text. A final sample of 85,003 participants had light data suitable for input to circadian rhythms models. Of this sample the majority of participants had 6 days of data remaining (65.8%), and there were 5 days in 21·2%, 4 days in 6·9%, and 3 days in 6·0%. No imputation was required in 27% of participants, and the remaining 73% had a median (IQR) of 0·57 (0.80) h of data imputed.

Initial conditions for Process P were defined according to each participant's weekly average midsleep time, commencing on the limit cycle. To minimize the transient effects of initial conditions on model outputs, we replicated each participant's data to 35 days in length (e.g., 7 days of light data were replicated 5 times, concatenated, and input to the model as a 35 day interval). Phase and amplitude were then calculated based on the last N days of the replicated times series, where N was number of days of data available for each participant.

### S11. Supplementary Analyses

#### Intra-Individual Stability of Light Exposure

We sought to determine whether participants' one-week light recordings were representative of their light environment across time. We examined light recordings in a subset of participants who completed up to four repeated actigraphy assessments across seasons (N = 2,988), occurring 3·23 ± 0·65 (M ± SD) years after initial assessment, with approximate three-month breaks between each. Lighting environments were highly stable within individuals across time, as indicated by an intra-class correlation coefficient of 0·803 (determined by a linear mixed model with crossed random effects structure including participant, assessment number, and time of day).

#### Mediation Analysis

Mediation was conducted using a generalized framework based on marginal structural models[4]. This approach was applied to estimate the direct and indirect effects of sleep duration on mortality. Total effect was estimated by including sleep duration as a predictor in an individual Cox regression. The mediating pathway from sleep duration (predictor) to light exposure (mediator) was assessed using linear regression, and mediation models were implemented using Cox regression. All models were adjusted for age, sex, ethnicity, physical activity, employment, income, deprivation, social activities, social visits, smoking status, urbanicity, and shift work status.

We assessed whether relationships of sleep duration with mortality was mediated by light exposure at night, since short sleep duration is known to correlate with mortality[5-6], and short or inefficient sleep may also correlate with light exposure at night. Sleep duration was split into short (<6 h) and normal sleepers (≥6 h). Light exposure at night was transformed using log(lux + 1) and included as a continuous predictor. There was a significant mediating effect of light at night on the sleep duration/mortality relationship: total effect: HR=1.33[1.18-1.49], p<.001; direct effect: HR=1.27[1.12-1.44], p<.001; indirect effect: HR=1.04[1.00-1.09], p=.04, where direct effect represents sleep/mortality relationship after accounting for the mediating effect of light, indirect effect represents the sleep/mortality relationship explained by light, and total effect represents the sleep/mortality relationship in the absence of light as a mediating variable.
