## Supplementary material for "Light at night and modeled circadian disruption predict higher risk of mortality: A prospective study in >88,000 participants": STROBE Checklist

STROBE Statement—Checklist of items that should be included in reports of ***cohort studies***

| | Item No | Recommendation | Manuscript Section |
|---|---|---|---|
| **Title and abstract** | 1 | (*a*) Indicate the study's design with a commonly used term in the title or the abstract | • Title |
| | | (*b*) Provide in the abstract an informative and balanced summary of what was done and what was found | • Abstract |
| **Introduction** | | | |
| Background/rationale | 2 | Explain the scientific background and rationale for the investigation being reported | • Introduction, Paragraphs 1-2 |
| Objectives | 3 | State specific objectives, including any prespecified hypotheses | • Introduction, Paragraph 3 |
| **Methods** | | | |
| Study design | 4 | Present key elements of study design early in the paper | • Introduction, Paragraph 3<br>• Methods: Overview |
| Setting | 5 | Describe the setting, locations, and relevant dates, including periods of recruitment, exposure, follow-up, and data collection | • Abstract<br>• Methods: Overview<br>• Results: Descriptive statistics<br>• Supplementary methods |
| Participants | 6 | (*a*) Give the eligibility criteria, and the sources and methods of selection of participants. Describe methods of follow-up | • Methods: Overview<br>• Supplementary methods |
| | | (*b*) For matched studies, give matching criteria and number of exposed and unexposed | • N/A |
| Variables | 7 | Clearly define all outcomes, exposures, predictors, potential confounders, and effect modifiers. Give diagnostic criteria, if applicable | • Methods: Overview<br>• Methods: Light exposure profiles<br>• Methods: Covariates |

| | | | |
|---|---|---|---|
| | | | • Supplementary methods |
| Data sources/ measurement | 8* | For each variable of interest, give sources of data and details of methods of assessment (measurement). Describe comparability of assessment methods if there is more than one group | • Methods: Overview<br>• Methods: Covariates<br>• Supplementary methods |
| Bias | 9 | Describe any efforts to address potential sources of bias | • Methods: Statistical analysis<br>• Supplementary methods |
| Study size | 10 | Explain how the study size was arrived at | • N/A |
| Quantitative variables | 11 | Explain how quantitative variables were handled in the analyses. If applicable, describe which groupings were chosen and why | • Methods: Statistical analysis<br>• Supplementary methods |
| Statistical methods | 12 | (*a*) Describe all statistical methods, including those used to control for confounding | • Methods: Statistical analysis<br>• Supplementary methods<br>• Supplementary analyses |
| | | (*b*) Describe any methods used to examine subgroups and interactions | • Methods: Statistical analysis<br>• Supplementary S7-9 |
| | | (*c*) Explain how missing data were addressed | • Methods: Light exposure profiles<br>• Supplementary methods |
| | | (*d*) If applicable, explain how loss to follow-up was addressed | • N/A |
| | | (*e*) Describe any sensitivity analyses | • Methods: Statistical analysis<br>• Supplementary S7-9 |
| **Results** | | | |
| Participants | 13* | (a) Report numbers of individuals at each stage of study—eg numbers potentially eligible, examined for eligibility, confirmed eligible, included in the study, completing follow-up, and analysed | • Methods: Overview<br>• Supplementary methods |
| | | (b) Give reasons for non-participation at each stage | • Methods: Overview |

| | | | |
|---|---|---|---|
| | | (c) Consider use of a flow diagram | • N/A |
| Descriptive data | 14* | (a) Give characteristics of study participants (eg demographic, clinical, social) and information on exposures and potential confounders | • Results: Descriptive statistics<br>• Results: Table 1 |
| | | (b) Indicate number of participants with missing data for each variable of interest | • Supplementary methods<br>• Results: Table 2 |
| | | (c) Summarise follow-up time (eg, average and total amount) | • Abstract<br>• Results: Descriptive statistics |
| Outcome data | 15* | Report numbers of outcome events or summary measures over time | • Results: Figure 1 |
| Main results | 16 | (*a*) Give unadjusted estimates and, if applicable, confounder-adjusted estimates and their precision (eg, 95% confidence interval). Make clear which confounders were adjusted for and why they were included | • Results: Tables 2-3<br>• Supplementary S7-9<br>• Methods: Statistical analysis |
| | | (*b*) Report category boundaries when continuous variables were categorized | • Methods: Statistical analysis |
| | | (*c*) If relevant, consider translating estimates of relative risk into absolute risk for a meaningful time period | • N/A |
| Other analyses | 17 | Report other analyses done—eg analyses of subgroups and interactions, and sensitivity analyses | • Supplementary S7-9<br>• Supplementary analyses<br>• Methods: Statistical analysis |
| **Discussion** | | | |
| Key results | 18 | Summarise key results with reference to study objectives | • Discussion: Paragraphs 1 |
| Limitations | 19 | Discuss limitations of the study, taking into account sources of potential bias or imprecision. Discuss both direction and magnitude of any potential bias | • Discussion: Paragraph 6 |

| | | | |
|---|---|---|---|
| Interpretation | 20 | Give a cautious overall interpretation of results considering objectives, limitations, multiplicity of analyses, results from similar studies, and other relevant evidence | • Discussion: Paragraphs 2-5, 7 |
| Generalisability | 21 | Discuss the generalisability (external validity) of the study results | • Discussion: Paragraph 6 |
| **Other information** | | | |
| Funding | 22 | Give the source of funding and the role of the funders for the present study and, if applicable, for the original study on which the present article is based | • N/A |
